# Prevalence and Associated Factors of Self-reported Domestic Elder Abuse among Rural Older Persons in Vietnam

**DOI:** 10.64898/2026.09.23.26363828

**Authors:** Tien Ngoc Thuy Doan, Van Hai Thi Hoang, Thanh Long Giang, Xiangming Fang, Dennis Reidy, Le Minh Giang

## Abstract

This study examined the prevalence of domestic elder abuse and examined factors in a province with a high aging index in Vietnam in 2025. Elder abuse was measured using the validated Vietnamese Domestic Elder Abuse Scale, covering psychological abuse, physical abuse, financial exploitation, neglect/abandonment. Logistic regressions identified factors associated with abuse. 29.44% of participants reported at least one abuse subtypes, with psychological abuse being the most common subtype (24.73%) and 9.82% experienced polyvictimization. Higher educational attainment was consistently associated with lower odds of all abuse subtypes. Having ≥4 chronic conditions was associated with higher odds of psychological abuse compared with having none (OR= 2.79, 95%CI: 1.29-6.00). Difficulties in daily basic activities were associated with physical abuse (OR=5.42, 95%CI: 1.80-16.36), financial exploitation (OR=3.66, 95%CI: 1.34-10.01), and neglect/abandonment (OR=4.09, 95%CI: 1.72-9.70) compared with no difficulties. Findings highlight the need for stronger policy implementation and lifelong learning to prevent elder abuse in Vietnam.

## Introduction

Globally, roughly one in six older persons (aged 60 and above) experienced some forms of elder abuse in community settings, although the true prevalence may be higher due to underreporting.^1–3^ Elder abuse is defined as inappropriate single or repeated acts within any trusting relationships which causes harm or distress to an older person.^4^ Forms of elder abuse vary depending on the victim-perpetrator relationships, with partners/spouses often commit psychological and physical abuse, children/grandchildren are more likely to perpetrate neglect/abandonment, and both family members and acquaintance commonly involve in financial exploitation.^5–11^

Domestic elder abuse is an emerging public health concern occurring across diverse settings and cultures. The urgency of this issue is amplified by rapid global population aging, with the number of older persons projected to more than double between 2015 and 2050.^12^ Rapid population aging, particularly in low-and middle income countries such as Vietnam, may intensify caregiving demands while the current health and social care systems for older population remain underdeveloped. Given that context, economic pressures on households and society, limited support resources, and increasing age-related physical and mental health impairments among older persons may heighten the risk of abusive behaviors within the homes. These challenges underscore the urgent need to prevent and address domestic elder abuse in Vietnam.

Recent empirical evidence from two waves of nationally representative surveys of older persons in Vietnam (OP&SHI 2019 and VNAS 2022) indicated that elder abuse is a critical domain influencing healthy aging status among the Vietnamese older population.^13^ Previous international studies also evidenced that elder abuse negatively affected their quality of life and overall well-being, thereby hindering the achievement of healthy aging in aging societies.^14,15^ From a policy perspective, tackling domestic elder abuse has become a crucial priority for the World Health Organization (WHO) and the United Nations (UN) under the Decade of Healthy Aging (2021-2030).^16^ The Government of Vietnam has shown a strong commitment by promulgating laws and regulations to emphasize the rights of older persons and combat the issue of domestic violence.^17–19^ Although its prevalence has been reported globally, elder abuse is likely underreported in Vietnam due to cultural factors. Older persons may feel ashamed to disclose domestic abuse and may fear legal consequences for the perpetrators, who are often family members or caregivers. Updated data on the past-year prevalence of elder abuse overall and by subtype in Vietnam remain limited. Hence, this study is our attempt to address two objectives: i) to estimate the past-year prevalence of domestic elder abuse overall and by subtypes in Ninh Binh province, Vietnam, and ii) to identify factors associated with each subtype of abuse. Ninh Binh province is a rural setting with the highest aging index in the Red River Delta region, making it a particularly relevant context for investigating this issue.^20^

In this study, domestic elder abuse was defined as experiencing at least one of the following subtypes: psychological abuse, physical abuse, financial exploitation, or neglect/abandonment among older persons. Physical abuse refers to acts that inflict physical pain or injury on an older person. Psychological abuse refers to verbal assaults, threats, intimidation, humiliation, or other behaviors that cause emotional distress. Financial exploitation involves the illegal or misuse or withholding of an older person’s financial resources. Neglect/abandonment refer to situations in which caregivers or responsible family members fail to provide necessary care, support, or assistance to an older person who depends on them for basic daily needs.^21^

In this study, the term “past-year” refers to the 12-month period preceding the survey, as data collection was conducted in late 2025. Accordingly, all prevalence estimates reported as past-year prevalence reflect experiences of elder abuse occurring within this 12-month recall period.

## Materials and methods

### Conceptual framework

We applied the social-ecological model of violence proposed by WHO,^4^ which conceptualizes elder abuse as the result of interacting factors across multiple levels. According to WHO (2002), there are four levels: individual level (including older persons and caregiver levels), interpersonal level (i.e. relationship between older persons and caregivers/family), community level, and societal level.^4^ Due to data availability, our study merely focused on factors at the individual and interpersonal levels, in which individual-level factors included age, gender, education, health conditions, functional limitations, while interpersonal-level factors included living arrangements, marital status, main source of income, and personal savings. This framework provides a theoretical basis for examining protective and risk factors of domestic elder abuse within the household context. The study framework is visualized in Figure 1.

**Figure 1:**
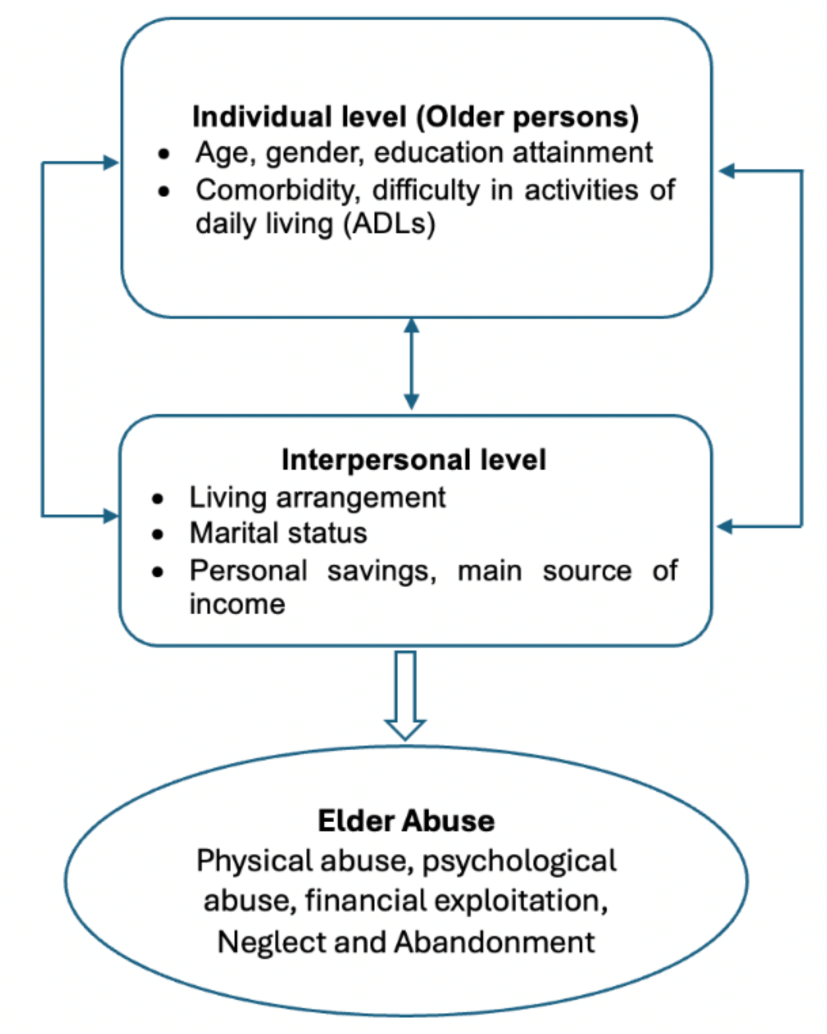
Study framework. Source: Own illustration.

### Setting and participants

According to the Article 2, Law on the Elderly 2009, older persons in Vietnam are defined as individuals aged fully 60 years and over (i.e., 61 years and above).^22^ Therefore, we conducted a cross-sectional study consisted of older persons aged 61 years and above in Ninh Binh province, Vietnam, which had the highest aging index in the Red River Delta region.^20^

The study sample were selected using a cluster random sampling method. Following WHO’s guidance for community-based surveys of noncommunicable disease risk factors, later extended to violence research, a design effect of 1.5 was applied.^23^ The sample size was calculated from the following formula:

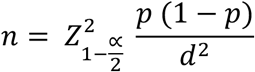

where:

n: the minimum required sample size;

Z_1-α/2_ = 1.96 (significant level α = 0.05; confident interval 95%);

d: standard error, d = 0.05;

p: the proportion of Vietnamese older persons suffering from abuse in community setting, which was derived from Dang et al. (2023) (31.67%).^24^

The initial sample size was 333. After applying a design effect of 1.5, the required sample was approximately 500; allowing for 20% non-response yielded a final minimum sample of 600.

Eligible participants were individuals aged 61 years and above, who had resided in the community in Ninh Binh province for at least 12 months prior to the survey and were able to communicate with interviewers. Older persons living in long-term care institutions or nursing homes, and those diagnosed with severe mental disorders or dementia by medical professionals, were excluded.

We applied a multi-stage cluster sampling combined with purposive selection of areas approach to select eligible participants. In the first sampling stage, we purposively selected Binh My commune, Ninh Binh province (which is the administrative merger of Binh My town, La Son commune, and Don Xa commune of former Ha Nam province). Subsequently, using simple random sampling, one village was randomly selected from each of the former communes/town. Then, from each selected village/residential cluster, 200 older persons were randomly selected at the household level within the community, yielding 600 eligible participants. With a minimum required sample size of 600, our study successfully recruited a total of 744 participants.

### Data collection

Face-to-face interviews were conducted using an administered questionnaire. Due to the sensitive nature of elder abuse and to protect potential abusive victims, all interviews were conducted in private settings. Family members, caregivers, or any other individuals were not allowed to be present during the interviews. Selected participants received telephone appointments beforehand to confirm private arrangements. This approach ensured participants feel safe and their confidentiality is maintained. Data were collected and stored using the data entry panel created in REDCap.

### Survey instrument

An administered questionnaire consisted of three components, including: i) demographic and socioeconomic, ii) health-related, and iii) elder abuse measure.

#### Domestic Elder Abuse Scale

Elder abuse was measured using the Domestic Elder Abuse Scale (DEAS), a structured instrument recently validated for use in the Vietnamese context (validation study under review), hereafter referred to as the Vietnamese version of Domestic Elder Abuse Scale.^25^ The original scale was developed by Heravi-Karimooi, et al. (2010) and comprised 49 items covering eight subtypes of elder abuse.^26^ The DEAS has been widely applied to measure the elder abuse across different settings through several validated versions.^27–29^ In this study, we adopted the Chinese-validated version of the DEAS because of the cultural similarities between China and Vietnam.^30^

The English version of the Chinese-validated DEAS was translated into Vietnamese and then back-translated into English to ensure preservation of the original meaning during the translation process. All feedback and revisions made during translation and refinement were fully documented by the research team. The finalized Vietnamese version was named the Vietnam–Domestic Elder Abuse Scale (Vie-DEAS).

Subsequently, the research team conducted a pilot test with 30 older persons, equivalent to 5% of the minimum required sample size. This step was conducted for the language, social context and cultural adaptations to the study population while preserving the original meaning and measurement properties of the scale. Following validation, four subtypes of abuse were retained: psychological abuse (8 items), physical abuse (4 items), financial exploitation (6 items), and neglect/abandonment (7 items). In the Vietnamese version, neglect/abandonment were combined into a single subtype. All items were rated on a 5-point Likert scale (never, occasionally, usually, always), with higher scores indicating a higher frequency of experiencing elder abuse. The instrument demonstrated good internal consistency in the present study, with Cronbach’s α ranging from 0.84 to 0.90 for four subtypes.^25^

In this study, the dependent variables were defined as binary outcomes capturing the experience of elder abuse in the past 12 months. Specifically, four binary outcomes were created to indicate whether older persons experienced each subtype of elder abuse (psychological abuse, physical abuse, financial exploitation, and neglect/abandonment). For each subtype, participants were considered exposed if they reported experiencing any abusive behavior at least “occasionally”, “usually”, or “always” on any corresponding item; those who answered “never” to all items were classified as not exposed. Accordingly, each subtype variable was coded as 0 (no experience) or 1 (experienced).

Additionally, the level of elder abuse victimization was defined as a categorical outcome based on the number of abuse subtypes experienced in the past year, including three groups: no elder abuse = 0, single-type elder abuse = 1, and elder abuse polyvictimization (experience of two or more subtypes) = 2.

#### Health-related characteristics

The health-related variables included i) comorbidity and ii) activity of daily living (ADL) limitation. In order to assess comorbidity conditions, older persons were asked whether they were diagnosed by health professionals with any of following chronic diseases, including hypertension, musculoskeletal diseases (e.g. arthritis, osteoarthritis), type 2 diabetes, chronic obstructive pulmonary disease, cardiovascular diseases, cancer, chronic kidney disease, chronic liver diseases (e.g., fatty liver), dyslipidemia (high blood lipids), stroke, depression, and dementia/Alzheimer’s disease. A *comorbidity* variable was created by summing the total number of self-reported conditions for each older person. This variable was categorized into four groups (0 diseases = 0; 1 disease = 1; 2–3 diseases = 3; and ≥4 diseases = 4), reflecting increasing levels of multimorbidity.

ADL limitation was used to determine whether older persons had difficulties in performing and accomplishing any basic activities (i.e. bathing, dressing, toileting, transferring, eating, and managing continence). The ADL limitation variable was dichotomized into two groups: No ADL limitations = 0 (no difficulty in performing all basic activities) and at least one ADL limitation = 1 (difficulty in performing at least one basic activity).

#### Demographic and socioeconomic characteristics

The demographic and socioeconomic characteristics included: age (continuous variable, measured in years); gender (male = 0; female = 1); living arrangement (living alone = 0; living with a spouse = 1; living with children only = 2; living with a spouse and other family members or caregivers = 3); marital status (never married = 0; current married = 1; widow/divorced/separate = 2); the main sources of income (current job = 0; pension = 1; social assistance = 2; saving and support from family members and relatives = 3); personal savings (No = 1; Yes = 2); and the highest educational attainment (incomplete primary school/illiteracy = 1; completed primary school = 2; completed lower secondary school =3; completed upper secondary school and above = 4).

All other categorical variables were presented with value 0 as reference groups.

### Data analyses

Descriptive analyses were presented as frequencies and percentages for categorical variables, as median and interquartile range (IQR) for continuous variables. In order to determine factors associated with elder abuse, we conducted logistic regression analyses for each subtype of elder abuse (i.e. psychological abuse, physical abuse, financial exploitation, and neglect/abandonment). In detail, bivariate logistic regression analyses were initially performed to examine the association between each independent variable and the outcomes. Variables with p-values lower than 0.15 in the bivariate analyses were considered including in the multivariate logistic regressions. Selection of variables for the multivariable analyses was further guided by theoretical relevance while considering the number of outcome events to minimize the risk of overfitting. We subsequently conducted multivariate logistic regressions to examine associated factors of elder abuse subtypes.

After examining factors associated with each subtype of elder abuse, a multivariable ordinal logistic regression model was conducted to investigate factors associated with increasing levels of victimization (i.e. no abuse, single-type abuse, and polyvictimization).

Adjusted odds ratios (aORs) and 95% confidence intervals (CIs) were reported. The p<0.05 was considered statistically significant level. We also reported p-value between 0.05 and 0.15, indicating marginal evidence of association.

### Ethical consideration

This study obtained ethical approval from the Institutional Review Board at Hanoi Medical University (Approval No. 2034/GCN-HMUIRB) and received permission from the People’s Committee of Binh My Commune for implementing this study. Eligible participants were asked to voluntarily participate in face-to-face interviews by signing in the inform consents before enrolling in the study. The written consent forms will be kept in locked cabinets. We explained clearly the research purposes, potential risks, and benefits (i.e. incentives) so respondents can decide to participate or withdraw at any time. All interviews were conducted in private spaces, without interruptions from family members, caregivers, or any other individuals. Any information gathered from respondents remained confidential and anonymous.

Regarding referral plan, we provided respondents a supporting documentation on how to deal with abuse situations. We provided them contacts to report abuse, including the hotlines for emergency assistance (i.e. 113 for police and 115 for emergency) and the national hotline for domestic violence prevention and control (111). Moreover, we encouraged victims or suspected victims to self-report their cases to People’s Committee, local police of the commune, ward or town, and/or the local Vietnam Association of the Elderly (VAE) where the elder abuse cases occurred.

## Results

Table 1 provides general information on older participants, as well as their characteristics stratified by exposure to elder abuse. Overall, the median age of older participants was 70.5 years (IQR: 65.5-76), the majority were female (66.26%), currently married (71.51%), and living with a spouse only (46.24%). Similar patterns were observed between older persons who experienced at least one form of abuse and those who did not; however, these differences were not statistically significant.

**Table 1.**
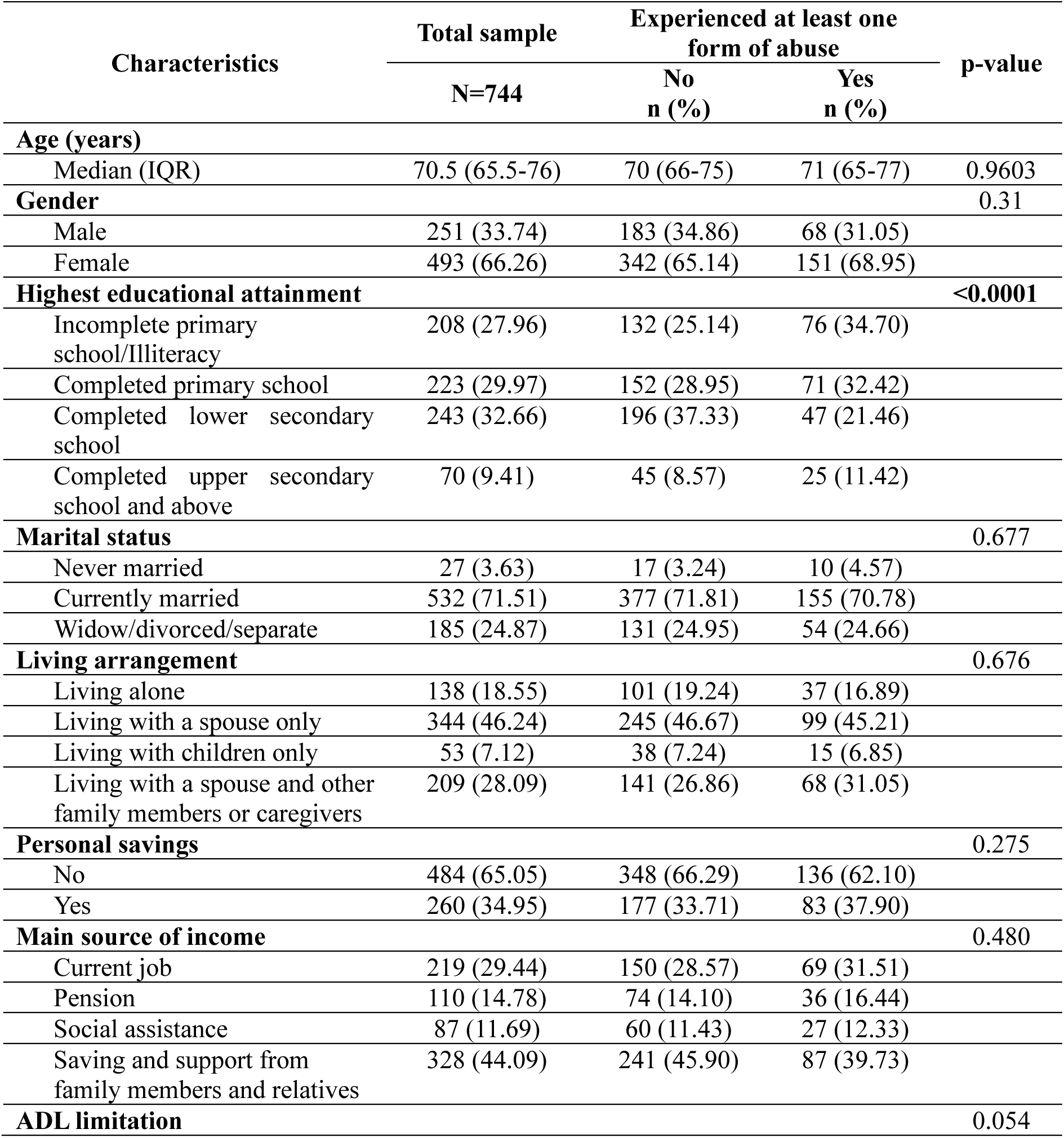
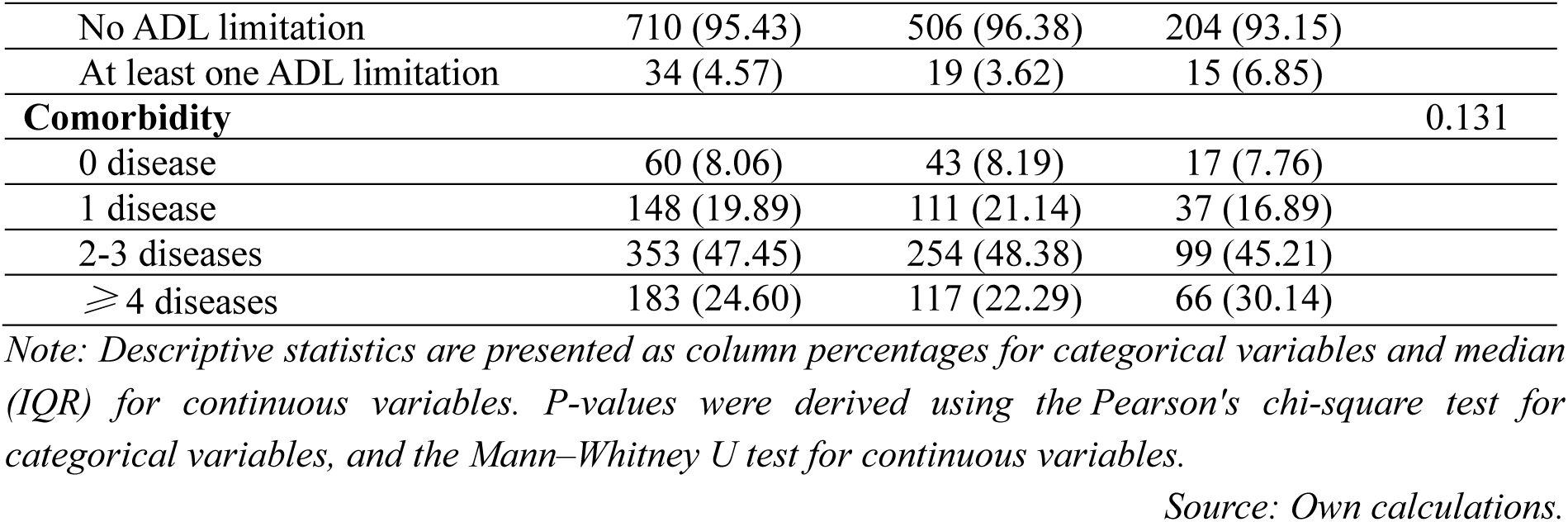
Descriptive characteristics of older participants by the exposure to elder abuse.

Regarding education, the largest proportion of participants in the overall sample had completed lower secondary school (32.66%), followed by those who completed primary school level. A similar pattern was observed among older persons who did not experience any form of abuse, with the majority completed lower secondary school (37.33%), followed by those who completed primary school (28.95%). However, this pattern differed among those who experienced at least one form of abuse. In details, the largest proportion of abused participants did not complete primary school or were illiterate (34.70%). These differences were statistically significant.

In terms of economic conditions, most of older persons had no savings (65.05%), and savings and financial support from family members and relatives were the main source of income (44.09%). Similar patterns were observed between older persons who experienced abuse and those who did not; however, these differences were not statistically significant.

Regarding health-related conditions, 95.43% of older participants found no ADL limitation. Moreover, the majority reported that they were diagnosed with two or more chronic diseases (71.85%). Similar patterns were observed in the abused and non-abused groups; with no statistically significant differences between them.

Regarding experiences of domestic elder abuse, 29.44% of older participants reported experiencing at least one form of abuse in the past 12 months. Among the subtypes, psychological abuse was the most prevalent (24.73%, n=184), followed by neglect/abandonment (7.12%, n=53), financial exploitation (5.38%, n=40), and physical abuse (4.03%, n=30) (Figure 1). These subtypes are not mutually exclusive. ngmethodsIn terms of the level of victimization, 19.62% experienced single-type elder abuse, while 9.82% experienced elder abuse polyvictimization in the past 12 months (Figure 1).

**Figure 1:**
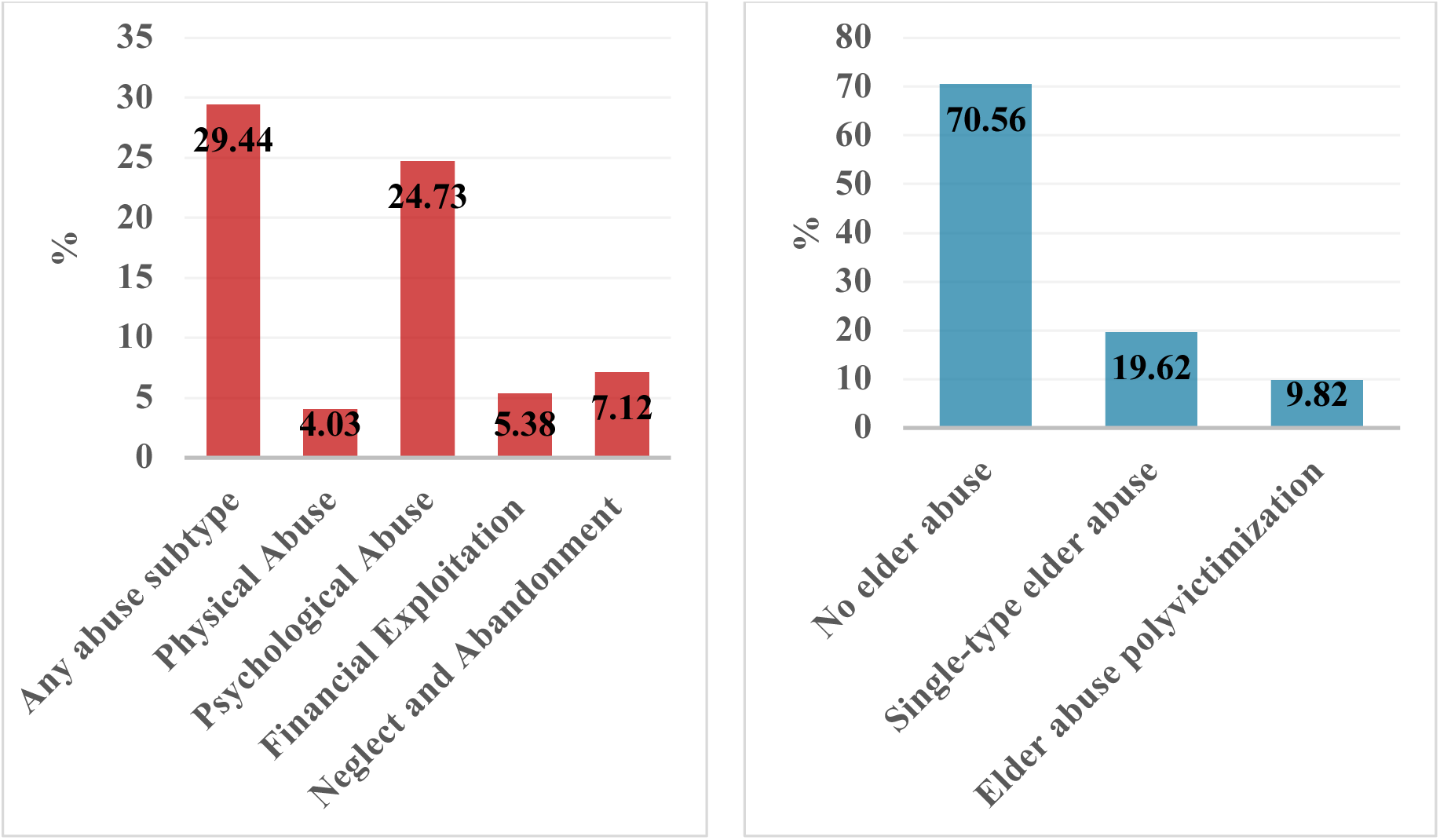
Past-year prevalence by abuse subtypes (left) and level of victimization (right) among rural older persons in Vietnam (%) Source: Own calculations.

Table 2 presents the results of univariate logistic regression analyses examining factors associated with subtypes of elder abuse, including physical abuse, psychological abuse, financial exploitation, neglect/abandonment. Regarding physical abuse, females had higher odds compared to males (OR=2.09, p=0.112), although this association was not statistically significant. Higher educational attainment was associated with lower odds of physical abuse, particularly among those who completed lower secondary education (OR=0.21, p=0.007) and upper secondary school and above (OR=0.18, p=0.107). Older persons who were widowed, divorced, or separated had lower odds to experience physical abuse compared to those never married (OR=0.22, p=0.048). Notably, older participants with at least one ADL limitation had significantly higher odds of experiencing physical abuse compared to those who did not (OR=4.72, p=0.003).

**Table 2.**
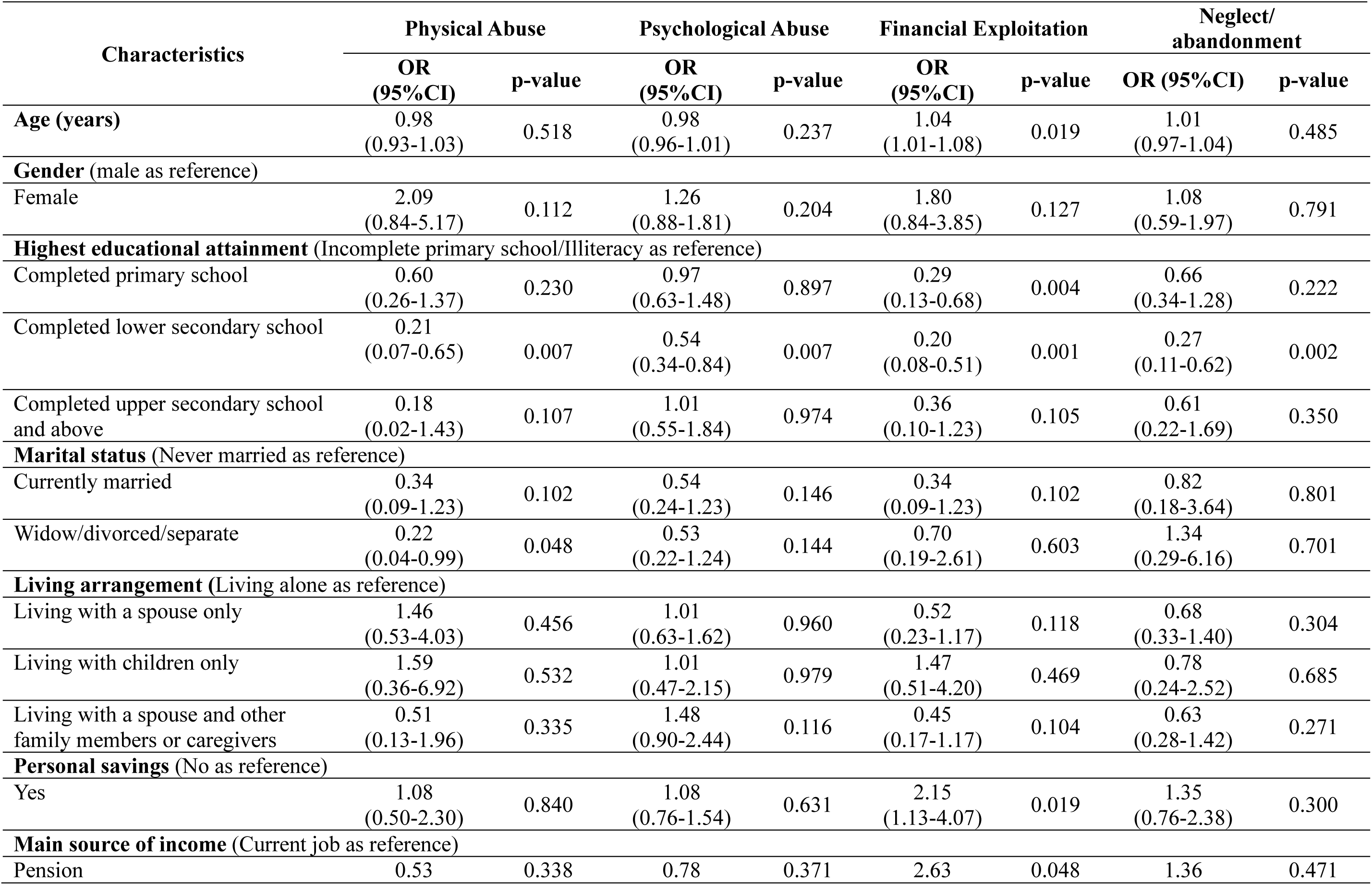
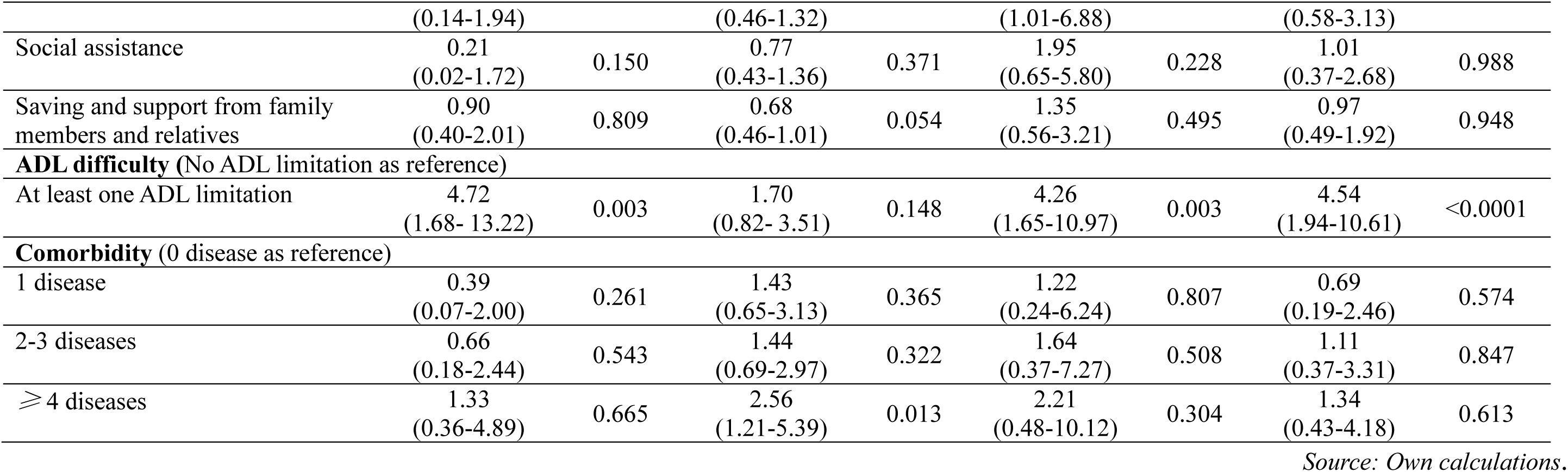
Univariable logistic regressions of factor associated with elder abuse subtypes among rural older adults in Vietnam.

As for psychological abuse, older participants being diagnosed with comorbidities (4 diseases and above) had significantly higher odds (OR=2.56, p=0.013). Those with at least one ADL limitation (OR = 1.70, p = 0.148) and those living with a spouse and other family members or caregivers (OR=1.48, p=0.116) appeared to have higher odds of experiencing psychological abuse, although these associations were not statistically significant. In contrast, older persons attaining lower secondary education was associated with reduced odds of experiencing psychological abuse (OR=0.54, p=0.007). Those who were currently married or widow/divorced/separate were less likely to suffer from psychological abuse; however, these associations were not statistically significant. Older participants relying on saving and financial support from family members was associated with reduced odds of experiencing psychological abuse (OR=0.68, p=0.054), although it did not reach conventional statistical significance.

In terms of financial exploitation, increasing age was significantly associated with higher odds (OR=1.04, p=0.019). Females had higher odds of experiencing financial exploitation (OR=1.80, p=0.127); however, it was not statistically significant. Older persons who attained higher educational levels were associated with lower odds of experiencing financial exploitation, particularly at primary (OR=0.29, p=0.004), lower secondary levels (OR=0.20, p=0.001), and upper secondary levels and above (OR=0.36, p=0.105). Those who were currently married had lower odds of experiencing financial exploitation than those who were never married (OR=0.34, p=0.105). Older persons who lived with a spouse only (OR=0.52, p=0.118) or with a spouse and other family members or caregivers (OR=0.45, p=0.104) were associated with lower odds to experience financial exploitation compared to those who lived alone. In contrast, those who had personal savings (OR=2.15, p=0.019) and received pension as the main source of income (OR=2.63, p=0.048) were associated with higher odds to experience financial exploitation. Similar to other subtypes, older adults with at least one ADL limitation were strongly associated with financial exploitation (OR=4.26, p=0.003). Although several variables met the selection threshold (p<0.15) in the univariate analysis for financial exploitation, the number of events was limited (n=40). Therefore, to avoid model overfitting, only a subset of variables was included in the multivariable regression model later based on prior literature and theoretical relevance, including age, the highest educational attainment, personal savings, and ADL difficulty.

Neglect/abandonment were strongly associated with education and at least one ADL limitation. In details, older persons who found difficulties in performing ADLs had substantially higher odds to experience neglect/abandonment (OR=4.54, p<0.0001). Those who had higher educational attainment, particularly lower secondary education, was associated with lower odds (OR=0.27, p=0.002).

The results of multivariate logistic regressions analyses examining factors associated with subtypes of elder abuse are illustrated in Table 3. For physical abuse, higher educational attainment remained protective after adjustment. In detail, older persons who completed lower secondary education had significantly lower odds compared to those with incomplete primary education or illiteracy (aOR=0.21, 95% CI: 0.07–0.66). Those who completed upper secondary education or above showed marginal evidence of association with physical abuse (aOR=0.20, 95%CI: 0.02–1.61). Those who were widowed, divorced, or separated had significantly lower odds of experiencing physical abuse compared to those who were never married (aOR=0.18, 95% CI: 0.04–0.85). In contrast, the odds of experiencing physical abuse among older persons with at least one ADL limitation were significantly 5.42 times higher than among those with no ADL difficulty.

**Table 3.** Multivariable logistic regressions of factor associated with elder abuse subtypes among rural older adults in Vietnam.

| Characteristics | Physical Abuse |  | Psychological Abuse |  | Financial Exploitation |  | Neglect/abandonment |  |
| --- | --- | --- | --- | --- | --- | --- | --- | --- |
|  | aOR | 95%CI | aOR | 95%CI | aOR | 95%CI | aOR | 95%CI |
| **Age (years)** | - | - | - | - | 1.01 | (0.97-1.05) | - | - |
| **Gender** (male as reference) |  |  |  |  |  |  |  |  |
| Female | 1.81 | (0.69-4.76) | - | - | - | - | - | - |
| **Highest educational attainment** (Incomplete primary school/Illiteracy as reference) |  |  |  |  |  |  |  |  |
| Completed primary school | 0.60 | (0.25-1.41) | 0.90 | (0.58-1.41) | **0.31\*** | **(0.13-0.73)** | 0.70 | (0.36-1.37) |
| Completed lower secondary school | **0.21\*** | **(0.07-0.66)** | **0.46\*** | **(0.28-0.74)** | **0.21\*** | **(0.08-0.54)** | **0.29\*** | **(0.12-0.67)** |
| Completed upper secondary school and above | 0.20† | (0.02- 1.61) | 0.93 | (0.48-1.80) | 0.35† | (0.10-1.25) | 0.67 | (0.24- 1.87) |
| **Marital status** (Never married as reference) |  |  |  |  |  |  |  |  |
| Currently married | 0.59 | (0.15- 2.29) | 0.49 | (0.18-1.30) | - | - | - | - |
| Widow/divorced/separate | **0.18\*** | **(0.04-0.85)** | 0.51† | (0.21-1.26) | - | - | - | - |
| **Living arrangement** (Living alone as reference) |  |  |  |  |  |  |  |  |
| Living with a spouse only | - | - | 1.31 | (0.65-2.62) | - | - | - | - |
| Living with children only | - | - | 1.08 | (0.48-2.42) | - | - | - | - |
| Living with a spouse and other family members or caregivers | - | - | **1.97\*** | (1.06-3.66) | - | - | - | - |
| **Personal savings** (No as reference) |  |  |  |  |  |  |  |  |
| Yes | - | - | - | - | **2.49\*** | **(1.28-4.84)** | - | - |
| **Main source of income** (Current job as reference) |  |  |  |  |  |  |  |  |
| Pension | - | - | 0.66 | (0.37-1.16) | - | - | - | - |
| Social assistance | - | - | 0.63† | (0.34-1.16) | - | - | - | - |
| Saving and support from family members and relatives | - | - | **0.58\*** | **(0.38-0.88)** | - | - | - | - |
| **ADL difficulty** (No ADL limitation as reference) |  |  |  |  |  |  |  |  |
| At least one ADL limitation | **5.42\*** | **(1.80- 16.36)** | 1.63 | (0.76-3.52) | **3.66\*** | **(1.34-10.01)** | **4.09\*** | **(1.72-9.70)** |
| **Comorbidity** (0 disease as reference) |  |  |  |  |  |  |  |  |
| 1 disease | - | - | 1.43 | (0.65-3.15) | - | - | - | - |
| 2-3 diseases | - | - | 1.53 | (0.73-3.21) | - | - | - | - |
| ≥ 4 diseases | - | - | **2.79\*** | **(1.29-6.00)** | - | - | - | - |

Note: Multivariate logistic regression results (“–” indicates variables not included in the model). Figures in bold indicate coefficients with statistically significant (\* $p < 0.05$ ); † $0.05 \leq p < 0.15$ (marginal evidence of association).
Source: Own calculations.

Regarding psychological abuse, older persons who completed lower secondary education were associated with reduced odds compared to those who had incomplete primary school or were illiterate (aOR=0.46, 95% CI: 0.28–0.74). Those who lived with a spouse and other family members or caregivers were significantly 1.97 times higher odds of suffering from psychological abuse than those who lived alone. Older persons diagnosed with four or more diseases were significantly associated with increased odds of experiencing psychological abuse compared to those who were not diagnosed with any diseases (aOR=2.79, 95% CI: 1.29–6.00). In addition, those who relied on saving and financial support from family members were associated with lower odds of experiencing this subtype of abuse compared to those who relied on current job (aOR=0.58, 95% CI: 0.38–0.88). Older persons who were widowed, divorced, or separated and those receiving social assistance showed tendencies toward lower odds of psychological abuse.

In terms of financial exploitation, educational attainment consistently remained a strong protective factor. In detail, older persons who completed primary education (aOR=0.31, 95% CI: 0.13–0.73) and lower secondary education (aOR=0.21, 95% CI: 0.08–0.54) had significantly lower odds compared to those with incomplete primary school or illiteracy. In contrast, the odds of financial exploitation among older persons with personal savings were 2.49 times higher than those who did not have savings (aOR=2.49, 95% CI: 1.28–4.84). Older persons with at least one ADL limitation had significantly higher odds of financial exploitation than those with no ADL limitation (aOR=3.66, 95% CI: 1.34–10.01). Those who completed upper secondary education or above showed a tendency toward lower odds of financial exploitation.

Neglect/abandonment was significantly associated with both educational attainment and functional status. Those who completed lower secondary education had lower odds of neglect/abandonment compared to those who did not complete primary school or were illiterate (aOR=0.29, 95% CI: 0.12–0.67). In contrast, the odds of neglect/abandonment among those with at least one ADL limitation were 4.09 times higher than those no ADL difficulties (aOR=4.09, 95% CI: 1.72–9.70).

Overall, ADL difficulty consistently emerged as a strong factor associated with increased odds of domestic elder abuse, particularly for physical abuse, financial exploitation, neglect/abandonment. Moreover, completing higher educational attainment, particularly at the lower secondary level, was also consistently a protective factor for all subtypes of elder abuse. Several variables showed tendencies toward association, suggesting potential relationships that require further investigation.

Table 4 illustrates factors associated with higher levels of elder abuse victimization. Educational attainment remained a strong protective association. Compared with older persons who were illiterate or had not completed primary school, those who completed lower secondary school had significantly lower odds of experiencing higher levels of victimization (aOR=0.31, 95% CI: 0.19–0.51). Consistent with the subtype-specific analyses, functional health status emerged as an important risk factor for higher levels of elder abuse victimization. Older persons with at least one ADL limitation had higher odds of being in a higher level of victimization compared with those without difficulties (aOR=2.26, 95% CI: 1.11–4.59). Living with a spouse and other family members or caregivers (aOR=1.56, 95% CI: 0.86–2.83) and relying on savings or family support as the main income source (aOR=0.68, 95% CI: 0.45–1.03) showed marginal associations with victimization levels. Other socio-demographic and health variables were not statistically significant.

**Table 4:** Order logistic regressions of factor associated with increasing levels of elder abuse victimization among rural older persons in Vietnam.

| Characteristics | Level of victimization |  |
| --- | --- | --- |
|  | aOR | 95%CI |
| **Age (years)** | 0.98 | (0.95- 1.01) |
| **Gender** (male as reference) |  |  |
| Female | 1.06 | (0.72- 1.55) |
| **Highest educational attainment** (Incomplete primary school/Illiteracy as reference) |  |  |
| Completed primary school | 0.72 | (0.47-1.09) |
| Completed lower secondary school | **0.31\*** | (0.19-0.51) |
| Completed upper secondary school and above | 0.68 | (0.36-1.31) |
| **Marital status** (Never married as reference) |  |  |
| Currently married | 0.63 | (0.24-1.64) |
| Widow/divorced/separate | 0.67 | (0.27-1.64) |
| **Living arrangement** (Living alone as reference) |  |  |
| Living with a spouse only | 1.38 | (0.71-2.69) |
| Living with children only | 1.18 | (0.71-2.48) |
| Living with a spouse and other family members or caregivers | 1.56† | (0.86-2.83) |
| **Personal savings** (No as reference) |  |  |
| Yes | 1.31 | (0.93-1.83) |
| **Main source of income** (Current job as reference) |  |  |
| Pension | 1.02 | (0.57-1.82) |
| Social assistance | 0.93 | (0.51-1.72) |
| Saving and support from family members and relatives | 0.68† | (0.45-1.03) |
| **ADL difficulty** (No ADL limitation as reference) |  |  |
| At least one ADL limitation | **2.26\*** | (1.11- 4.59) |
| **Comorbidity** (0 disease as reference) |  |  |
| 1 disease | 0.77 | (0.39-1.52) |
| 2-3 diseases | 0.97 | (0.52-1.81) |
| ≥ 4 diseases | 1.47 | (0.77-2.82) |

*Note: Multivariable ordinal logistic regression results. Figures in bold indicate coefficients with statistically significant (\* $p < 0.05$ ); † $0.05 \leq p < 0.15$ (marginal evidence of association).*
*Source: Own calculations*

## Discussion

In this study, we found that the prevalence of domestic elder abuse was 29.44% in Binh My commune, Ninh Binh province, Vietnam. This estimate appears lower than those reported in studies from Iran (75.4%), Nepal (56.4%), and Bangladesh (71.5%),^27,31–33^ but higher than estimates from India (9.36%), China (10%), Japan (12.3%), and Australia (15%).^34–37^ However, direct comparisons should be interpreted with caution due to substantial differences in study design, definitions of elder abuse, measurement tools, and socio-cultural contexts across studies.

First, although these prevalence estimates were derived from community-based samples, they were measured using different scales to assess elder abuse. Some studies adopted measurements comprising a set of questionnaires, for instance the Hwalek-Sengstock Elder Abuse Screening Test and Vulnerability to Abuse Screening Scale,^38,39^ while others only used a single question to identify the abusive behaviors.^36^ Differences in measurement tools can lead to variations in definitions, the abusive behaviors, and the subtypes of abuse, all of which can significantly affect the observed prevalence.

Second, variations may also reflect differences in the scope of these studies, including sample size, geographic coverage, and the demographic characteristics of the populations studied. Studies with smaller samples may report higher prevalence compared with larger and multi-region studies, partly due to sampling variability. For instance, several studies were conducted in specific cities or provinces,^27,31,32^ whereas others were conducted as nationally representative surveys.^36,37^

Third, socio-cultural contexts play a critical role in shaping both the occurrence and the reporting of elder abuse. Differences in social norms, family dynamics, the perceived role of older persons, and sensitivity toward reporting abusive behaviors can contribute to observed differences in prevalence. For instance, in Southeast Asian cultures, admitting or reporting abuse may be considered shameful, viewed as a private matter that should remain within the family, which can lead to underreporting despite actual incidents.

In our study, regarding abuse subtypes, psychological abuse was the most prevalent form (24.73%). Other subtypes were also observed in our study sample; however, their prevalence was lower. More specifically, the prevalence of neglect/abandonment, financial exploitation, and physical abuse was 7.12%, 5.38%, and 4.03%, respectively. The prevalence and its factors associated with each subtype of elder abuse, as presented in the following section.

### Psychological and Physical abuse

Given the most common subtypes of abuse observed in this study, our findings are consistent with previous research.^28,36,40,41^ This consistency suggests that psychological abuse may be prevalent among older populations across diverse cultural and geographic contexts. Older persons may experience reduced social participation, increased economic dependency, and functional impairments, which can contribute to feelings of isolation and vulnerability.^42–44^ When family members or caregivers fail to meet their emotional or practical expectations, older persons may feel misunderstood or neglected, thereby exacerbating their sense of insecurity. Conversely, caregiving can be stressful and demanding tasks, and when caregivers experience burden or frustration, interpersonal tensions may escalate, increasing the risk of abusive behaviors, especially psychological and physical abuse.^45,46^ The interpersonal relationships between older persons and caregivers or family members should be further investigated to have a comprehensive understanding on this field.

Regarding psychological abuse in our study, older persons living with a spouse and other family members or caregivers were approximately twice as likely to experience psychological abuse compared with those living alone, further emphasizing the importance of interpersonal dynamics in shaping abuse risk. Moreover, older persons with four or more chronic diseases were 2.79 times more likely to experience psychological abuse than those without any diagnosed diseases. Taken together, the combined effects of comorbidity and co-residential living arrangements may reinforce the vulnerability of older persons within dependent and potentially strained caregiving relationships, as discussed above.

In terms of physical abuse, our study found that the past-year experience of this subtype was the least common among our sample. The estimated prevalence was lower than that reported in some previous studies.^28,40,46^ These direct comparisons should be interpreted with caution due to differences in the definitions and measurements of physical abuse across studies.^36,38,39^ Moreover, in many Southeast Asian cultural contexts, particularly in Vietnam, domestic abuse is often regarded as a private family matter, and disclosing such experiences may be associated with shame. As a result, older persons may refuse to admit or report incidents of abuse, leading to a potential underestimation of the true prevalence.

Our findings further revealed that older persons having at least one ADL limitation were 5.42 times more likely to experience physical abuse than those without such difficulties. This finding again highlights the role of functional dependency as an important risk factor for physical abuse.^45,46^ Older persons who require assistance with daily activities often rely heavily on family members or caregivers, which may increase caregiving burden and stress. Thus, it can lead to frustration, conflict, and a higher risk of abusive behaviors.

Interestingly, older persons who were widowed, divorced, or separated were less likely to experience physical abuse compared with those who had never married. One possible explanation is that older persons who have never married can be more likely to live with extended family members or depend on relatives for later life, which may increase the frequency of close interactions and potential conflicts. In contrast, widowed or previously married older persons may have greater autonomy, more social networks, or different living arrangements that reduce prolonged exposure to caregiving stress and interpersonal tensions within households.

### Financial exploitation

The misuse of older persons’ resources is recognized as a violation their legal rights. Financial exploitation is challenging to identify and measure because it often occurs in private, involves complex financial or legal arrangements between older persons and their family members or even acquaintance. Similar to other subtypes of abuse, it is often perceived as a sensitive family matter, which contributes to underreporting and difficulties in verification.

In our study, only 5.38% of older persons reported experiencing financial exploitation. This estimate is broadly comparable to findings from some settings, such as Australia (2.1%), India (4.2%).^37,38^ However, substantially higher prevalence has been reported in other contexts, including Bangladesh (60.4%) and Iran (35.4%).^28,31^ These wide variations should be interpreted with caution, as differences in study design, definitions of financial exploitation, measurement tools, sampling strategies, and sociocultural contexts may limit direct comparability across studies.

Older persons who had personal savings were 2.49 times more likely to experience financial exploitation than those without savings. In addition, functional impairment remained an important risk factor. Older persons with at least one ADL limitation were 3.66 times more likely to experience financial exploitation compared with those no ADL limitation. These findings suggest that older persons who were more dependent in daily functioning and who possessed personal savings may become more vulnerable to financial exploitation. The risk of financial exploitation is influenced by multiple factors. First, functional dependence increases reliance on family members or caregivers, especially for routine financial transactions, which may create opportunities for misuse or unauthorized control of financial resources. Previous studies also supported this finding.^47,48^ In addition, having personal savings may increase the perceived economic value of older persons, and when combine with functional or cognitive impairments, thereby increasing the risk of exploitation.

### Neglect/abandonment

Our finding was consistent with previous literature suggesting that neglect/abandonment was one of common subtypes of domestic elder abuse.^28,31,37^ As health deteriorates, older persons may become increasingly dependent on family members or caregivers for assistance with daily living. When caregiving demands exceed available resources, the risk of unmet needs and unintentional or intentional neglect may arise. Our findings support this explanation, as older persons with at least one ADL limitation were more than four times more likely to experience neglect/abandonment. In the Vietnamese context, caregiving is largely provided by family members, which places substantial pressure on their time and effort when caring for older persons. Moreover, limited caregiving knowledge and skills may further contribute to inadequate care and, in some cases, eventual abandonment.

### Level of elder abuse victimization

Reduction in physical health continued to be a key risk factor for higher levels of elder abuse victimization (i.e. sing-type abuse and elder abuse polyvictimization). Older persons at least one ADL limitation often require greater assistance with routine tasks, which may increase caregiving demands and the frequency of close interactions with family members and caregivers. Hence, older persons were more likely to experience higher levels of victimization, including single-type abuse and even polyvictimization (experiencing two or more abuse subtypes). In Vietnam, the formal support systems and resources for aged care remain limited, this increasing dependence may contribute to caregiver stress, frustration, and interpersonal conflict, thereby elevating the risk of multiple forms of abuse, including physical abuse, psychological abuse, financial exploitation, and neglect/abandonment.

Overall, our findings underscore the need for stronger protective policies and implementation from governmental and social organizations following the 4P approach (Prevention–Protection–Promotion–Provision of services). Expanding access to home-based care and developing caregiver support programs may reduce caregiving burden, prevent abuse and neglect, even prevent escalation from single-type abuse to polyvictimization. In details, providing caregiver incentives, such as financial subsidies, respite care services, or training programs, may help families sustain long-term caregiving and reduce stress and burnout. In addition, the provision of trained home-based care workers under a formal financing scheme (e.g., social health insurance), particularly for older persons with severe health conditions, could further reduce the burden on families and improve the quality and continuity of care.

Regarding higher educational attainment, this factor continued to play as a consistent protective factor against increased levels of elder abuse victimization. This finding, once again, highlights the importance of lifelong learning as a long-term protective resource across the life course. Education may enhance health literacy, communication skills, problem-solving ability, and awareness of personal rights. Therefore, education enables older persons to maintain greater autonomy, better recognize abusive behaviors, and seek for assistance. These capacities may reduce the likelihood of progressing from no abuse to single-type abuse, and further to polyvictimization. Moreover, strengthening financial protection mechanisms through education and promotion is particularly critical in the Vietnamese context. This includes improving financial literacy, enhancing awareness of common fraud and exploitation schemes, and strengthening community and institutional safeguards to protect older persons from repeated or multiple forms of financial abuse.

### Study strengths and limitations

To the best of our knowledge, this is the first study to examine the prevalence of elder abuse by specific subtypes in a rural setting in Vietnam, including psychological abuse, physical abuse, financial exploitation, neglect/abandonment, using the Vie-DEAS, which was recently validated in Vietnam.^25^ The Vie-DEAS provides detailed measurement of each subtype, whereas previous studies in Vietnam typically relied on a limited number of general questions capturing abuse in broader terms.^24,49,50^ Moreover, this is our attempt to estimate the prevalence of elder abuse polyvictimization. Given the limited evidence on elder abuse in Vietnam, it is crucial to conduct larger-scale studies building on these findings. Moreover, our study attempted to capture multiple factors associated with domestic elder abuse, including socio-economic and health status. These findings contribute to a more comprehensive understanding of the patterns and correlates of domestic elder abuse in rural Vietnamese settings.

Several limitations were acknowledged in this study. First, the cross-sectional design precludes causal inference, and the observed associations between elder abuse and its potential determinants should therefore be interpreted with caution. Second, elder abuse was measured using self-reported experiences over the previous 12 months, which may be subject to recall and information bias. Third, although the overall prevalence of abuse was not negligible, the number of cases within some subtypes was relatively small, resulting in wide confidence intervals and limited statistical power to detect associations in the multivariable analyses. Future investigations should consider longitudinal study designs to better understanding the causal relationships between risk factors and elder abuse. Moreover, larger-scale studies would help improve statistical power and provide more precise estimates. In addition, further studies investigating information on other sources, such as caregivers and community, may help reduce reporting bias and provide a more comprehensive understanding of domestic elder abuse in Vietnam.

## Conclusion

This study reported a past-year prevalence of domestic elder abuse of 29.44%, with 744 older participants indicating that they had experienced at least one subtype of abuse in 2025. Psychological abuse was the most prevalent subtype, affecting 24.73% of participants. Our findings also revealed 9.82% of older persons experienced two or more subtypes (elder abuse polyvictimization). Educational attainment and health status consistently emerged as protective and risk factors, respectively, across all subtypes and levels of victimization. Strengthening home-based care support could improve the quality of care for older persons while reducing caregiver stress and burnout. Promoting lifelong learning, including raising awareness of abusive behaviors and encouraging older adults to seek support when needed, is also essential.

## DECLARATIONS

### Ethics approval and consent to participate

Ethical approval from the Institutional Review Board at Hanoi Medical University (Approval No. 2034/GCN-HMUIRB). Written informed consent was obtained from all participants prior to participation.

### Availability of data and materials

The dataset is not publicly available due to data-sharing restrictions associated with the study funded under training grant D43TW012188.

### Authors’ contributions and agreement

All authors contributed to the study protocol and design. Data collection, manuscript drafting, and figures/tables preparation was done by Tien Ngoc Thuy Doan. Intellectual inputs for editing manuscript were provided by Long Thanh Giang, Van Thi Hai Hoang, Xiangming Fang, Dennis Reidy, and Le Minh Giang. Data collections and data analysis were provided by Tien Ngoc Thuy Doan. Manuscript review was done by Long Thanh Giang, Van Thi Hai Hoang, Xiangming Fang, Dennis Reidy, and Le Minh Giang. All authors read and approved the final version of the manuscript.

## Acknowledgements

This research was supported by the training grant D43TW012188, Consortium for Violence Prevention Research, Implementation, and Training for Excellence (CONVERGE), PI Yount & MPI Giang, from the Fogarty International Center of the National Institutes of Health. The authors are gratefully acknowledged by training grant D43TW012188 awarded for funding the conduct of this research. The authors also sincerely thank all participating older persons for providing the information necessary for this study.

